# Optic Nerve Lesion Volume, White Matter Hyperintensities, and Brain Volumetrics in Multiple Sclerosis: A Multi-Sequence MRI-Based Analysis

**DOI:** 10.1101/2025.04.25.25326426

**Authors:** Adrian Korbecki, Tomasz Konopczyński, Oktawian Hawro, Aleksandra Blachucik, Krzysztof Winiarczyk, Kamil Litwinowicz, Michał Sobański, Agata Zdanowicz-Ratajczyk, Justyna Korbecka, Justyna Chojdak-Łukasiewicz, Anna Pokryszko-Dragan, Joanna Bladowska, Anna Zimny

## Abstract

**Objectives:** To investigate the relationship between optic nerve lesion volume (ONLV), measured on double inversion recovery (DIR) MRI, and other radiological biomarkers of disease burden in multiple sclerosis (MS), including white matter hyperintensities (WMHs), T1-weighted hypointensities (“black holes”), and brain volumetrics. This study aims to determine whether ONLV correlates with a more severe neurodegenerative profile and could serve as a potential biomarker for disease severity.

**Materials and Methods:** In this cross-sectional study, 212 MS patients underwent 3T MRI including 3D T1W, FLAIR, and DIR sequences. Optic nerve lesions were manually segmented on DIR images and quantified volumetrically. Patients were grouped by optic nerve involvement: none (n = 59), unilateral (n = 60), or bilateral (n = 93). WMHs were segmented and anatomically categorized using an AI-based tool. T1W hypointensities were extracted via FreeSurfer, and brain volumetrics were assessed using a machine learning–based segmentation algorithm on both 3D T1W and FLAIR images. Statistical comparisons and correlation analyses were performed using multivariate models adjusted for total intracranial volume.

**Results:** ONLV was positively correlated with periventricular (r = 0.365, p < 0.001), deep white matter (r = 0.165, p = 0.005), and juxtacortical (r = 0.163, p = 0.007) WMHs. Patients with bilateral optic nerve involvement exhibited significantly higher WMH burden, greater T1W hypointensities (β = 10.91, p < 0.001), and more pronounced cerebral white matter atrophy (β = –107.02, p = 0.016). Regional brain atrophy was most evident in structures along the visual pathway (e.g., cuneus, fusiform gyrus, pericalcarine cortex, and thalamus). Additionally, periventricular WMH volume was significantly associated with global brain atrophy, including cortical and subcortical gray matter loss.

**Conclusions:** This study demonstrates that ONLV, as quantified on DIR MRI, is associated with increased lesion burden, irreversible white matter damage, and widespread brain atrophy in MS. These findings suggest that ONLV may serve as a potential imaging biomarker of disease severity and could be integrated into standard MRI protocols for MS assessment. Future longitudinal studies are warranted to validate ONLV as a marker of disease progression and treatment response.

## Introduction

Optic nerve involvement is a well-established feature of multiple sclerosis (MS), often manifesting as optic neuritis (ON), an inflammatory demyelinating event with potential long-term implications for visual function and neurodegeneration (Gabilondo et al., 2014). With the 2024 revision of the McDonald criteria incorporating optic nerve lesions as a fifth anatomical site for demonstrating dissemination in space (DIS), the role of optic nerve pathology in MS diagnosis and disease monitoring has gained further clinical significance. Although conventional imaging techniques such as double inversion recovery (DIR) and short tau inversion recovery (STIR) sequences facilitate the detection of optic nerve abnormalities, volumetric assessment of optic nerve lesions remains largely unexplored (Hadhoum et al., 2016; Hodel et al., 2014).

Previous studies have established a relationship between ON-related optic nerve damage and both anterograde and retrograde neurodegenerative processes within the central nervous system (CNS)(Calvi et al., 2022; Davion et al., 2023; Gabilondo et al., 2014). Anterograde degeneration, propagating from the optic nerve to the visual pathways and occipital cortex, has been correlated with cortical thinning and structural alterations in the posterior brain regions. Conversely, retrograde effects, extending from the CNS toward the optic nerve, may be driven by white matter pathology and global brain atrophy associated with MS progression (Gabilondo et al., 2014). Notably, MRI-based volumetric analyses have provided valuable insights into whole-brain and regional atrophy patterns in MS, yet little attention has been given to the quantitative volumetric assessment of lesions within the optic nerves.

In this study, we aimed to bridge this knowledge gap by comparing MS patients with optic nerve lesions detected on the DIR sequence against MS patients without such lesions. Importantly, this is the first study to measure the volume of optic nerve lesions in the DIR sequence, rather than relying solely on lesion presence or count. Our objective was to determine whether the presence and volume of optic nerve lesions are associated with a more severe radiological profile, including a higher burden of white matter hyperintensities on 3D fluid-attenuated inversion recovery (FLAIR), a greater presence of hypointensities (“black holes”) on T1-weighted images (T1W), and more extensive brain atrophy assessed from both 3D T1W and 3D FLAIR sequences. By systematically analyzing these radiological differences, we aimed to evaluate whether optic nerve lesion volume (ONLV) has the potential to serve as an indicator of greater disease burden and should be tested as a valuable biomarker for assessing disease severity and progression in MS patients in future longitudinal studies.

## Materials and methods

### Study population

This single-center study was carried out at the University Hospital of Wroclaw, Poland. The study cohort consisted of 212 patients diagnosed with MS according to the 2017 revision of the McDonald criteria by experienced neurologists. All patients were 18 years or older and were receiving disease-modifying therapy (DMT) appropriate for their MS subtype at the time of the study.

Of the 212 patients, 153 patients (72.17%) had optic nerve lesions detected on the DIR sequence, while 59 patients (27.83%) did not have optic nerve involvement, serving as the control group. Among those with optic nerve lesions, 93 patients (60.78%) had bilateral involvement, while 60 patients (39.22%) had unilateral involvement. The mean age of patients without optic nerve lesions was 40.97 years (range: 21–64 years, SD: 10.73), with 74.58% being female. In comparison, patients with optic nerve lesions had a mean age of 40.64 years (range: 21–68 years, SD: 11.67), with 73.2% being female.

The cohort primarily included individuals with relapsing-remitting MS (RRMS) (n = 194), while secondary progressive MS (SPMS) (n = 11) and primary progressive MS (PPMS) (n = 5) were less represented. The distribution of multiple sclerosis subtypes was relatively balanced between the groups. Each patient underwent a comprehensive neurological evaluation, including an assessment of the Expanded Disability Status Scale (EDSS) during clinical visit.

All participants provided written informed consent before inclusion in the study. The study protocol was approved by the local ethics committee and conducted in accordance with the Declaration of Helsinki.

### MRI image acquisition

All participants underwent brain MRI using a Philips Ingenia 3T system equipped with a 32-channel SENSE head coil. Standard patient positioning and coil setup were maintained to ensure uniform image acquisition across all subjects. The MRI protocol included a 3D T1W, 3D FLAIR sequence, and 3D DIR sequence, optimized for volumetric and lesion analysis.

### 1. 3D T1W acquisition

T1-weighted images were obtained using a three-dimensional gradient-echo sequence with the following parameters: repetition time (TR) = 7.9 ms, echo time (TE) = 3.5 ms, flip angle = 8°, slice thickness = 1 mm, matrix size = 252 × 250, and field of view (FOV) = 250 mm. The images were acquired in the axial orientation, ensuring high-resolution visualization of brain structures for volumetric assessment.

### 2. 3D FLAIR acquisition

Fluid-attenuated inversion recovery images were obtained in sagittal plane using a three-dimensional turbo spin-echo sequence with TR = 4800 ms, TE = 289.1 ms, inversion time (TI) = 1650 ms, flip angle = 90°, echo train length = 167, slice thickness = 1.12 mm, matrix size = 224 × 224, and FOV = 250 mm. The acquisition parameters provided high-resolution contrast for detecting white matter lesions and volumetric brain analysis.

### 3. 3D DIR acquisition

Double inversion recovery images were acquired in sagittal plane using a three-dimensional inversion recovery sequence to enhance lesion detection by simultaneously suppressing cerebrospinal fluid (CSF) and white matter signals. The parameters were TR = 5500 ms, TE = 287.6 ms, TI = 2550 ms, echo train length = 173, slice thickness = 1.3 mm, spacing between slices = 0.65 mm, matrix size = 208 × 208, and FOV = 250 mm, flip angle = 90°. The acquisition was optimized to enhance lesion conspicuity in the brain and optic nerves.

### Optic nerve volumetric analysis

The volumetric assessment of optic nerve lesions was performed using a manual segmentation approach in 3D Slicer. The primary imaging sequence for segmentation was 3D DIR, which enhances contrast by suppressing CSF and white matter signals, thereby improving the visualization of optic nerve abnormalities. Manual annotation of lesions within the optic nerves was initially conducted by a medical engineer in a slice-by-slice manner along the axial and coronal plane (**Fig.1**). To ensure segmentation accuracy, results were further refined and validated by an experienced neuroradiologist (at least 7 years of experience). In cases where segmentation uncertainty arose due to the artifacts, additional verification was performed by cross-referencing the DIR images with the patient’s FLAIR images.

**Figure 1.**
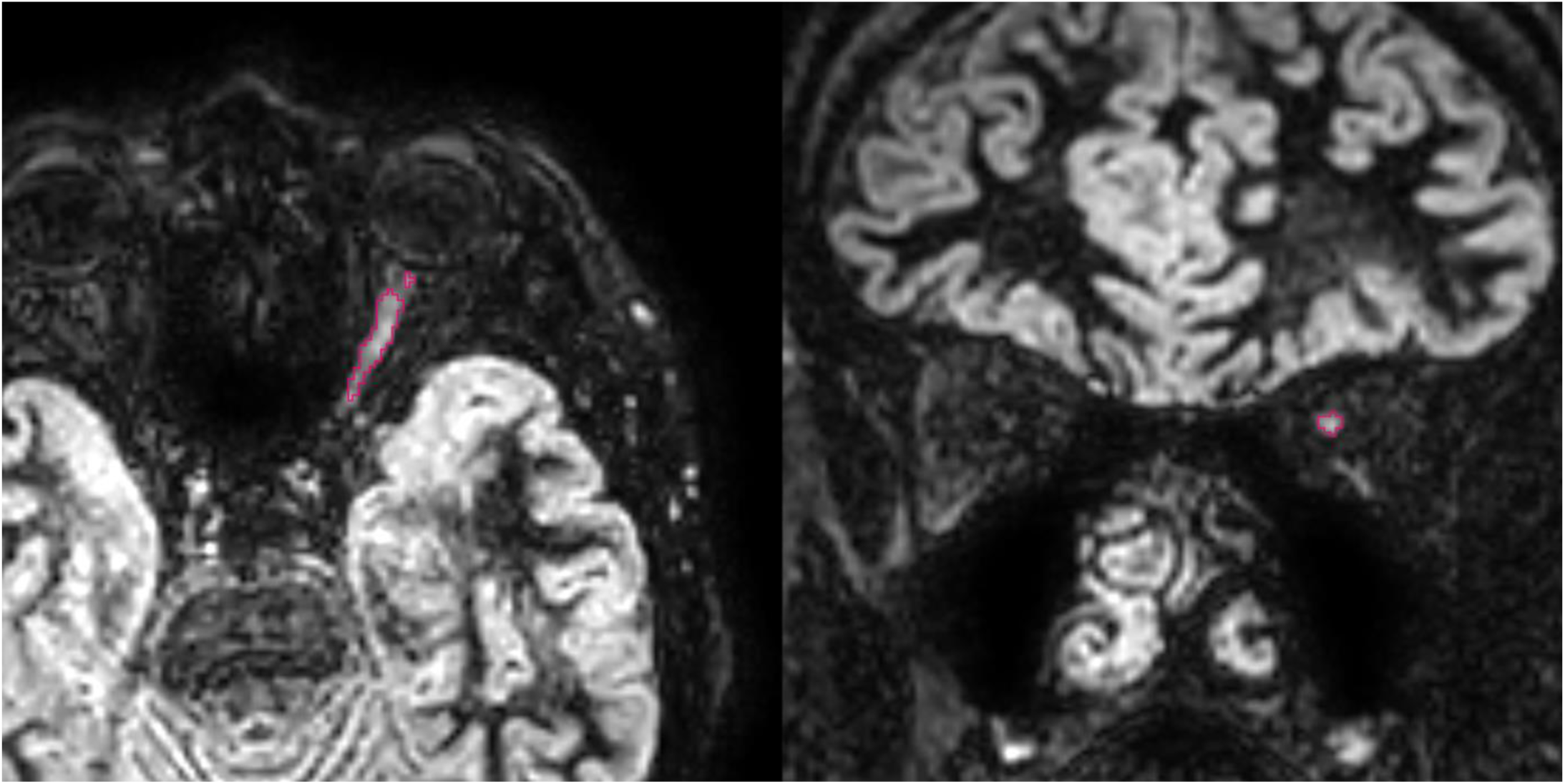
Detection of optic nerve lesions on DIR MRI sequence. Axial (left) and coronal (right) views of a DIR MRI sequence showing optic nerve lesions. The highlighted regions (marked in red) indicate areas of abnormal hyperintensity consistent with multiple sclerosis-related optic nerve involvement.

Following segmentation, volumetric calculations were performed to determine the total ONLV. Patients were categorized into three subgroups: no optic nerve involvement, unilateral involvement, and bilateral involvement, to facilitate comparative analysis across these groups.

### White matter hyperintensities segmentation – MS plaques

White matter hyperintensities (WMH) segmentation was conducted on the 3D FLAIR sequence using a proprietary software developed by Hetalox **(Fig. 2)**. The AI system comprises two custom-designed machine learning models that leverage adaptive 3D U-Nets - one for WMH segmentation and another for brain segmentation to ensure precise localization (Isensee et al., 2021). This AI-based method enables quantitative lesion assessment, including lesion burden, which represents the total volume of WMH in cubic centimeters, lesion count, and lesion load, which quantifies the percentage of total white matter volume affected by lesions.

**Figure 2.**
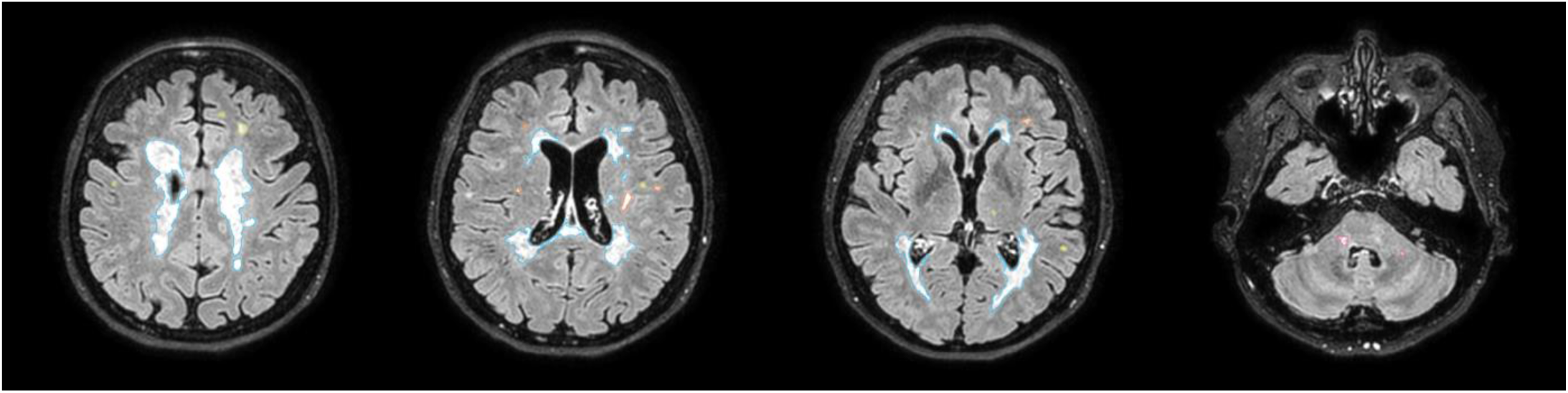
White matter lesions in multiple sclerosis on 3D FLAIR MRI. Axial slices from a 3D FLAIR MRI sequence demonstrating extensive WMHs in a patient with multiple sclerosis. Lesions are segmented and color-coded based on their anatomical location: periventricular lesions are outlined in blue, juxtacortical lesions in orange, deep white matter lesions in yellow, and infratentorial lesions in pink.

Additionally, the algorithm provides regional classification of WMH based on anatomical localization, distinguishing lesions as periventricular, deep white matter, juxtacortical, and infratentorial. All lesion-related data were compiled into a patient-specific report (**Table 1**).

**Table 1.** WMH burden volume metrics in an example MS patient.

| Category | Value |
| --- | --- |
| Total WMH Burden (volume in cm<sup>3</sup>) | 5.079 |
| Lesion Count | 41 |
| Lesion Load (%) | 0.2549 |
| **Periventricular WMH Burden (volume in cm<sup>3</sup>)** | **4.3728** |
| Periventricular Lesion Count | 5 |
| Periventricular Lesion Load (%) | 0.2194 |
| **Deep White Matter WMH Burden (volume in cm<sup>3</sup>)** | **0.2635** |
| Deep White Matter Lesion Count | 12 |
| Deep White Matter Lesion Load (%) | 0.0132 |
| **Juxtacortical WMH Burden (volume in cm<sup>3</sup>)** | **0.4058** |
| Juxtacortical Lesion Count | 21 |
| Juxtacortical Lesion Load (%) | 0.0204 |
| **Infratentorial Lesion Burden (cm<sup>3</sup>)** | **0.0372** |
| Infratentorial Lesion Count | 3 |
| Infratentorial Lesion Load (%) | 0.0019 |

The AI-based segmentation can be applied to both 3D T1W and 3D FLAIR sequences or using FLAIR alone. In this study, both 3D T1W and 3D FLAIR images were utilized as they were included in the standardized scanning protocol. This combined approach allows for a more comprehensive evaluation of lesion distribution and volumetric characteristics.

### Black holes segmentation - T1W hypointensities

In addition to WMH segmentation, we performed automated segmentation of T1-weighted hypointensities, commonly referred to as black holes, using FreeSurfer (Fischl, 2012). This software enables automatic detection of hypointense lesions within the white matter, providing volumetric information on lesion burden. The segmentation process quantified the number of hypointense lesions as well as the total volume of T1-weighted hypointensities in cubic millimeters. This approach allowed for a comprehensive assessment of chronic tissue damage and neurodegeneration, as black holes are associated with irreversible axonal loss in MS pathology.

### Brain volumetric assessment

The volumetric assessment of brain structures was conducted separately on 3D T1W and 3D FLAIR sequences using Hetalox’s software (**Fig.3**). The software employs a machine learning algorithm for brain structure segmentation, trained using the SynthSeg approach. By leveraging extensive synthetic data generation during training, it ensures robust and accurate segmentation of brain scans with any contrast and resolution, accommodating diverse acquisition protocols and scanner types (Billot et al., 2023). This AI-based tool employs a segmentation approach comparable to FreeSurfer’s Sequence Adaptive Multimodal SEGmentation (SAMSEG), facilitating robust, sequence-adaptive segmentation of brain structures without the need for extensive preprocessing (Puonti et al., 2016).

**Figure 3.**
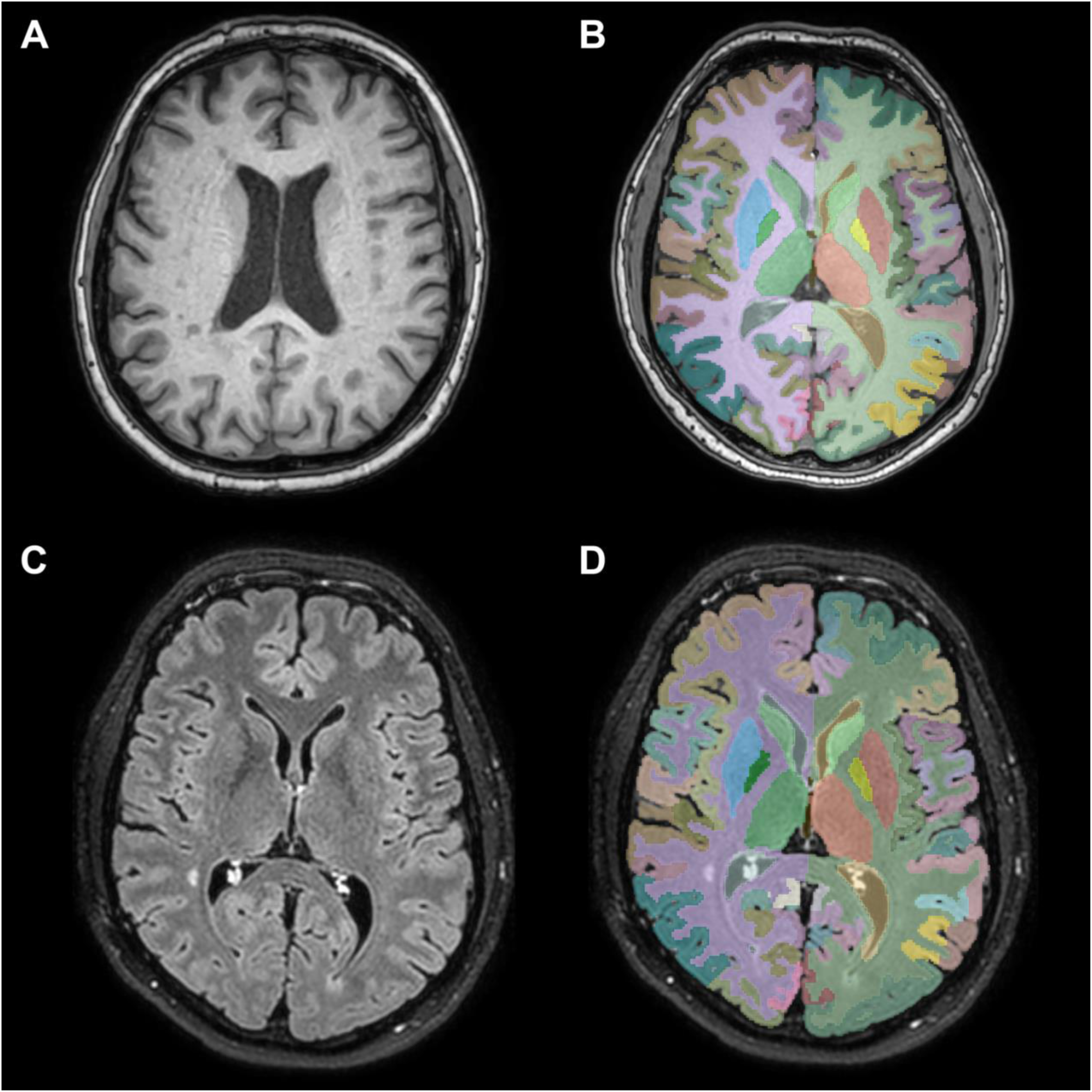
Brain MRI segmentation of cortical and subcortical structures. Representative axial slices demonstrating brain segmentation using structural MRI. (A) T1-weighted axial MRI scan. (B) Automated cortical and subcortical segmentation overlaid on the T1-weighted scan. (C) 3D FLAIR axial scan. (D) Automated segmentation applied to the 3D FLAIR scan, enabling regional volumetric analysis.

The Hetalox algorithm performs whole-brain segmentation, delineating 101 distinct brain structures, including both cortical and subcortical regions. Cortical parcellation was based on the Desikan-Killiany atlas, which provides a standardized anatomical framework for regional volume analysis. This segmentation methodology allows for an accurate volumetric assessment of brain structures in MS patients, considering both global brain volume and regional atrophy patterns.

To ensure consistency, the volumetric segmentation was applied to both 3D T1W and 3D FLAIR sequences, as both were included in the standard scanning protocol. Additionally, volumetric measurements obtained from the two sequences were systematically compared to assess potential differences and to enhance the precision of the volumetric analysis. This comparative approach aimed to exploit volume estimations across different MRI contrasts, ensuring robustness in structural assessments. A detailed list of the segmented brain structures is provided in **Table 2**.

**Table 2:** Segmented brain structures in 3D T1W and 3D FLAIR sequences with example values.

| Index | Structure | Vol. (cm3) | Index | Structure | Vol. (cm3) | Index | Structure | Vol. (cm3) |
| --- | --- | --- | --- | --- | --- | --- | --- | --- |
| 1 | Total Intracranial Volume | 1605.498 | 35 | Left Caudal Anterior Cingulate | 3.048 | 69 | Right Caudal Anterior Cingulate | 3.615 |
| 2 | Left Cerebral White Matter | 237.959 | 36 | Left Caudal Middle Frontal Gyrus | 6.346 | 70 | Right Caudal Middle Frontal Gyrus | 6.925 |
| 3 | Left Cerebral Cortex | 254.967 | 37 | Left Cuneus | 4.336 | 71 | Right Cuneus | 3.654 |
| 4 | Left Lateral Ventricle | 10.989 | 38 | Left Entorhinal Cortex | 3.577 | 72 | Right Entorhinal Cortex | 3.915 |
| 5 | Left Inferior Lateral Ventricle | 0.586 | 39 | Left Fusiform Gyrus | 10.196 | 73 | Right Fusiform Gyrus | 10.599 |
| 6 | Left Cerebellum White Matter | 16.671 | 40 | Left Inferior Parietal Lobule | 11.732 | 74 | Right Inferior Parietal Lobule | 14.479 |
| 7 | Left Cerebellum Cortex | 55.547 | 41 | Left Inferior Temporal Gyrus | 9.596 | 75 | Right Inferior Temporal Gyrus | 9.938 |
| 8 | Left Thalamus | 6.675 | 42 | Left Isthmus of Cingulate Cortex | 2.512 | 76 | Right Isthmus of Cingulate Cortex | 3.913 |
| 9 | Left Caudate | 4.038 | 43 | Left Lateral Occipital Cortex | 10.597 | 77 | Right Lateral Occipital Cortex | 10.024 |
| 10 | Left Putamen | 5.999 | 44 | Left Lateral Orbitofrontal Cortex | 6.796 | 78 | Right Lateral Orbitofrontal Cortex | 7.484 |
| 11 | Left Pallidum | 1.762 | 45 | Left Lingual Gyrus | 7.612 | 79 | Right Lingual Gyrus | 8.754 |
| 12 | Third Ventricle | 1.287 | 46 | Left Medial Orbitofrontal Cortex | 5.547 | 80 | Right Medial Orbitofrontal Cortex | 5.844 |
| 13 | Fourth Ventricle | 1.622 | 47 | Left Middle Temporal Gyrus | 9.034 | 81 | Right Middle Temporal Gyrus | 9.880 |
| 14 | Brain Stem | 22.442 | 48 | Left Parahippocampal Gyrus | 2.651 | 82 | Right Parahippocampal Gyrus | 3.444 |
| 15 | Left Hippocampus | 4.356 | 49 | Left Paracentral Lobule | 4.819 | 83 | Right Paracentral Lobule | 4.660 |
| 16 | Left Amygdala | 1.866 | 50 | Left Pars Opercularis | 4.442 | 84 | Right Pars Opercularis | 3.716 |
| 17 | Cerebrospinal Fluid (CSF) | 363.365 | 51 | Left Pars Orbitalis | 1.845 | 85 | Right Pars Orbitalis | 4.380 |
| 18 | Left Accumbens Area | 0.702 | 52 | Left Pars Triangularis | 4.647 | 86 | Right Pars Triangularis | 5.494 |
| 19 | Left Ventral Diencephalon | 4.208 | 53 | Left Pericalcarine Cortex | 3.355 | 87 | Right Pericalcarine Cortex | 3.070 |
| 20 | Right Cerebral White Matter | 239.043 | 54 | Left Postcentral Gyrus | 10.084 | 88 | Right Postcentral Gyrus | 11.959 |
| 21 | Right Cerebral Cortex | 258.539 | 55 | Left Posterior Cingulate Cortex | 5.474 | 89 | Right Posterior Cingulate Cortex | 4.606 |
| 22 | Right Lateral Ventricle | 9.604 | 56 | Left Precentral Gyrus | 12.724 | 90 | Right Precentral Gyrus | 12.809 |
| 23 | Right Inferior Lateral Ventricle | 0.511 | 57 | Left Precuneus | 8.190 | 91 | Right Precuneus | 9.753 |
| 24 | Right Cerebellum White Matter | 16.930 | 58 | Left Rostral Anterior Cingulate Cortex | 2.800 | 92 | Right Rostral Anterior Cingulate Cortex | 2.420 |
| 25 | Right Cerebellum Cortex | 56.434 | 59 | Left Rostral Middle Frontal Gyrus | 13.541 | 93 | Right Rostral Middle Frontal Gyrus | 14.650 |
| 26 | Right Thalamus | 6.313 | 60 | Left Superior Frontal Gyrus | 18.624 | 94 | Right Superior Frontal Gyrus | 20.232 |
| 27 | Right Caudate | 4.044 | 61 | Left Superior Parietal Lobule | 10.564 | 95 | Right Superior Parietal Lobule | 11.830 |
| 28 | Right Putamen | 5.951 | 62 | Left Superior Temporal Gyrus | 8.073 | 96 | Right Superior Temporal Gyrus | 8.821 |
| 29 | Right Pallidum | 1.712 | 63 | Left Supramarginal Gyrus | 8.550 | 97 | Right Supramarginal Gyrus | 9.626 |
| 30 | Right Hippocampus | 4.420 | 64 | Left Frontal Pole | 2.668 | 98 | Right Frontal Pole | 3.195 |
| **31** | Right Amygdala | 1.841 | **65** | Left Temporal Pole | 3.702 | **99** | Right Temporal Pole | 3.578 |
| **32** | Right Accumbens Area | 0.683 | **66** | Left Transverse Temporal Gyrus | 1.644 | **100** | Right Transverse Temporal Gyrus | 2.265 |
| **33** | Right Ventral Diencephalon | 4.432 | **67** | Left Insula | 6.209 | **101** | Right Insula | 6.486 |
| **34** | Left Banks of the Superior Temporal Sulcus | 2.540 | **68** | Right Banks of the Superior Temporal Sulcus | 2.911 |  |  |  |

## Statistical analysis

Statistical analyses were conducted using R version 4.3.3. To compare demographic variables between groups, normality was assessed with the Shapiro–Wilk test. Because age deviated from normality, it was compared across the three groups with the Kruskal–Wallis test, followed by pairwise Wilcoxon rank-sum tests. Sex distribution was analysed with Pearson’s χ² test. Multivariate linear regression models were used to evaluate the impact of regional WMHs (periventricular, deep white matter, juxtacortical, and infratentorial) and optic nerve involvement assessed on DIR images on brain volumetric measures, including total brain volume, subcortical gray matter, cortical gray matter, total gray matter, cerebral white matter, and volumes of individual cortical and subcortical structures. To control for individual differences in head size, all models were adjusted for total intracranial volume (TIV). Multiple testing was controlled using the Benjamini-Hochberg procedure. ANOVA was conducted with Tukey’s HSD post hoc test to examine the impact of optic nerve involvement on T1 hypointensities and plaque burden.

## Results

Patients with two affected nerves were significantly older than those with one (42.6 vs 37.5 years), and no other demographic variables differed significantly between groups.

The analysis demonstrated a direct correlation between ONLV and higher volumes of periventricular (r = 0.365, p < 0.001), deep white matter (r = 0.165, p = 0.005), and juxtacortical (r = 0.163, p = 0.007) WMH lesions measured on 3D FLAIR. However, no correlation was found between ONLV and infratentorial lesions (**Fig. 4**). Summary results for WMHs across different localizations, grouped by the number of affected optic nerves, are presented in **Table 3**.

**Figure 4.**
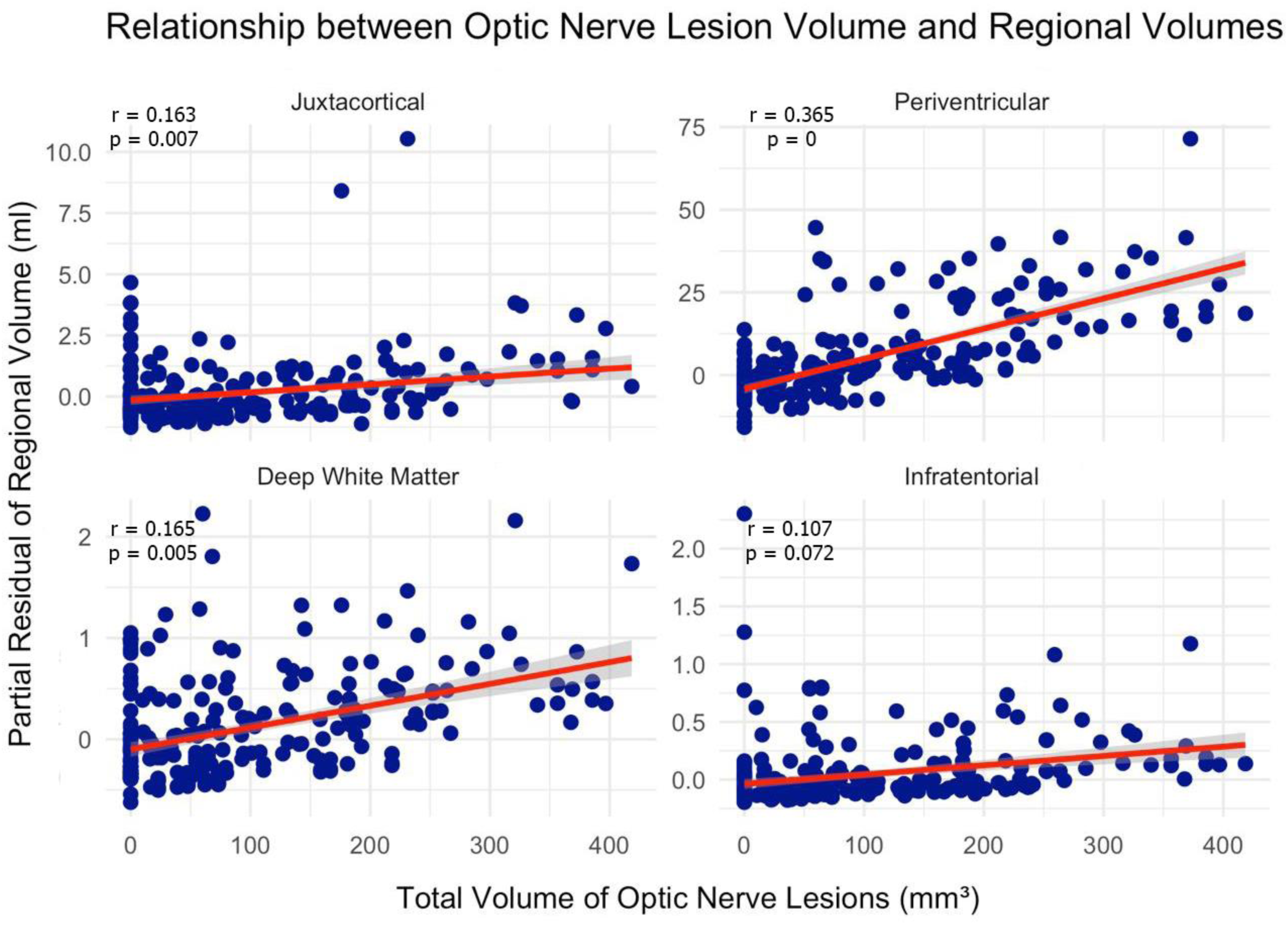
Linear regression between total volume of optic nerve lesions and regional white matter lesion volume. Scatterplots show significant positive correlations between optic nerve lesion volume and WMHs volume in deep white matter, juxtacortical, and periventricular regions.

**Table 3.** Regional white matter hyperintensities, white matter hypointensities, and brain volume metrics in MS patients by number of optic nerves involved.

| Variables | Number of nerves involved | Range | Mean ± SD | Median (IQR) |
| --- | --- | --- | --- | --- |
| **White matter hyperintensities (ml)** |  |  |  |  |
| Total WMH burden | 0 | 1.71–29.42 | $9.22 \pm 5.96$ | 6.23 (10.47) |
| | 1 | 0.81–42.47 | $6.07 \pm 6.63$ | 4.39 (3.42) |
| | 2 | 1.03–70.35 | $20.60 \pm 13.97$ | 19.29 (20.34) |
| Periventricular WMH | 0 | 1.30–24.89 | $7.21 \pm 5.09$ | 5.98 (8.39) |
| | 1 | 0.66–40.92 | $5.25 \pm 5.99$ | 4.20 (2.59) |
| | 2 | 0.63–64.51 | $18.49 \pm 13.18$ | 16.26 (19.87) |
| Deep White Matter WMH | 0 | 0.01–1.61 | $0.44 \pm 0.48$ | 0.31 (0.39) |
| | 1 | 0.00–2.71 | $0.25 \pm 0.41$ | 0.10 (0.20) |
| | 2 | 0.00–2.42 | $0.66 \pm 0.40$ | 0.58 (0.32) |
| Juxtacortical WMH | 0 | 0.00–5.29 | $1.45 \pm 1.44$ | 0.83 (1.68) |
| | 1 | 0.00–3.29 | $0.41 \pm 0.61$ | 0.17 (0.20) |
| | 2 | 0.00–11.18 | $1.23 \pm 1.42$ | 0.97 (1.07) |
| Infratentorial WMH | 0 | 0.00–2.50 | $0.13 \pm 0.29$ | 0.06 (0.18) |
| | 1 | 0.00–0.92 | $0.16 \pm 0.29$ | 0.02 (0.13) |
| | 2 | 0.00–1.20 | $0.22 \pm 0.24$ | 0.16 (0.32) |
| **White matter hypointensities (mm<sup>3</sup>)** |  |  |  |  |
| | 0 | 445.90–10287.50 | $1849.57 \pm 1863.68$ | 1288.50 (1306.17) |
| | 1 | 428.60–14689.70 | $1901.69 \pm 2535.65$ | 1208.70 (909.80) |
| | 2 | 459.60–19589.40 | $4275.95 \pm 3830.99$ | 2714.40 (4426.70) |
| **Brain volume metrics (mm<sup>3</sup>)** |  |  |  |  |
| Brain volume | 0 | 924855.00–1472364.00 | $1106976.12 \pm 142699.21$ | 1081978.00 (292464.00) |
| | 1 | 790025.00–1333435.00 | $1159595.26 \pm 124689.95$ | 1174129.00 (219686.00) |
| | 2 | 784158.00–1447141.00 | $1045517.90 \pm 104038.00$ | 1049535.00 (116571.00) |
| Subcortical gray matter volume | 0 | 44382.00–72126.00 | $57229.29 \pm 7004.66$ | 56738.00 (8769.00) |
| | 1 | 34151.00–65783.00 | $58821.30 \pm 5423.76$ | 60723.00 (9491.00) |
| | 2 | 35428.00–73038.00 | $51321.77 \pm 5757.54$ | 50930.00 (7848.00) |
| Cortical gray matter volume | 0 | 388537.26–599881.71 | $462224.27 \pm 51027.11$ | 465985.08 (103026.78) |
|  | 1 | 334876.12–548765.43 | 482664.10 ± 48042.72 | 500325.55 (89541.47) |
|  | 2 | 346033.10–659128.02 | 447005.77 ± 43766.04 | 447313.23 (49756.96) |
| Total gray matter volume | 0 | 534471.74–819613.71 | 629861.08 ± 65082.02 | 626362.63 (121954.78) |
|  | 1 | 469548.12–736770.43 | 654477.24 ± 62121.00 | 678875.51 (131160.28) |
|  | 2 | 482031.10–857768.02 | 606543.41 ± 55995.79 | 605855.82 (60868.88) |
| Cerebral white matter volume | 0 | 336027.00–624669.00 | 453588.97 ± 77002.60 | 436368.00 (159056.00) |
|  | 1 | 301643.00–568261.00 | 479534.19 ± 63848.24 | 468677.00 (98435.00) |
|  | 2 | 256775.00–559137.00 | 416527.67 ± 53646.02 | 417822.00 (54655.00) |

Additionally, it was noted that patients with bilateral optic nerve involvement exhibited a significantly higher total plaque burden compared to those with no optic nerve involvement (p < 0.001). Interestingly, patients with unilateral optic nerve involvement did not exhibit a statistically significant difference in total plaque burden compared to those without optic nerve lesions (**Fig. 5**).

**Figure 5.**
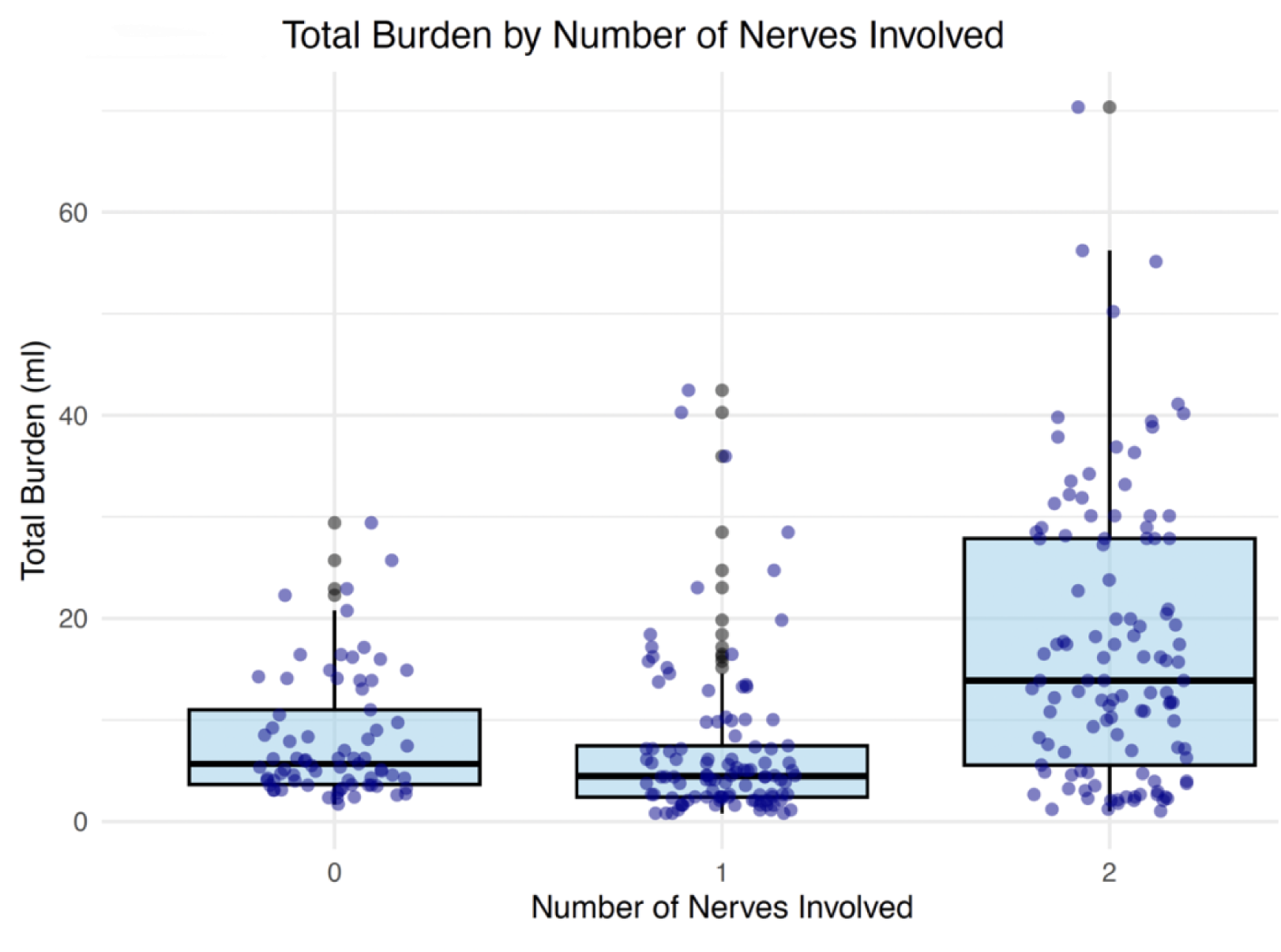
Total white matter lesion burden by number of optic nerves involved. This boxplot illustrates the distribution of total white matter lesion burden (in milliliters) across three patient groups categorized by the number of optic nerves involved: no involvement (0), unilateral involvement (1), and bilateral involvement (2).

The analysis of T1W hypointensities (black holes) revealed that patients with bilateral optic nerve lesions exhibited a significantly higher total volume of T1-weighted hypointensities within the white matter compared to those with either unilateral or no optic nerve lesions. Furthermore, when comparing the volumes of T1W hypointensities with the ONLV, a direct positive correlation was observed (p < 0.001; **Fig. 6**). A breakdown of T1W hypointensities volumes stratified by the number of affected optic nerves is also provided in **Table 3**.

**Figure 6.**
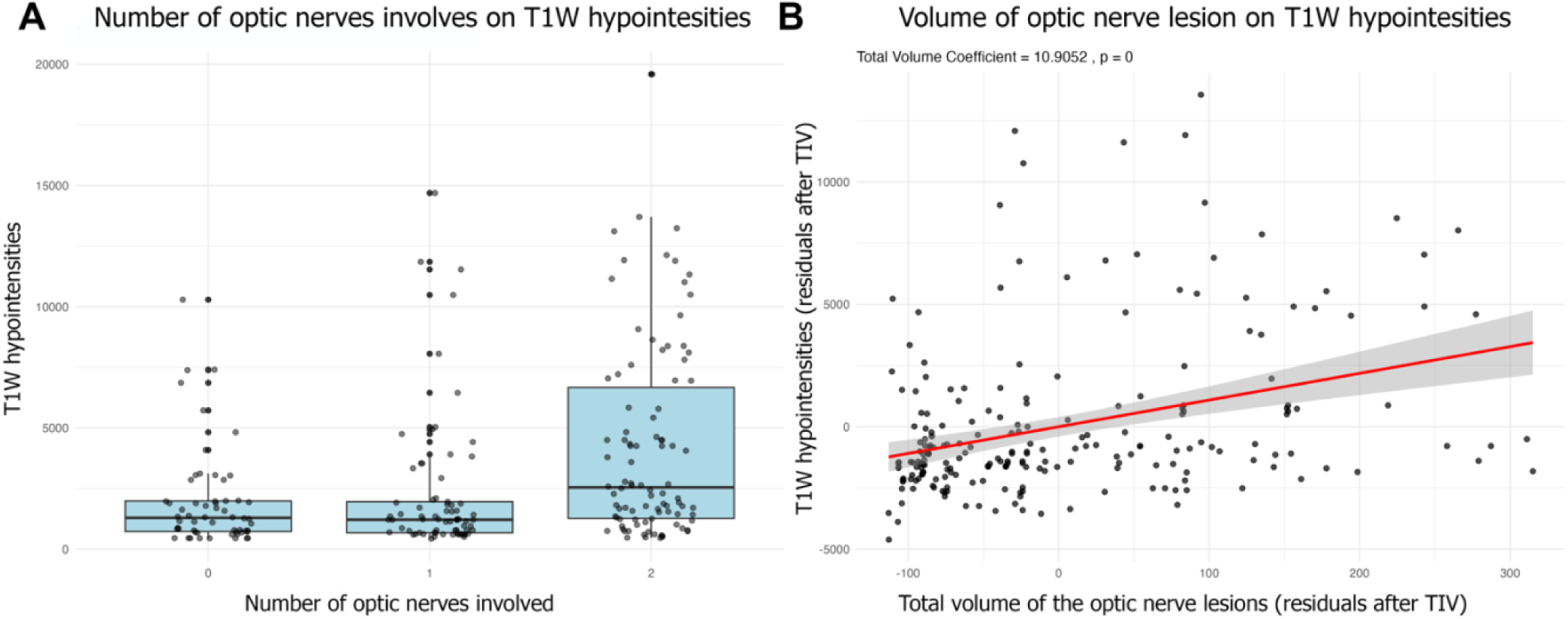
Relationship between optic nerve involvement and T1-weighted hypointensities (black holes). (A) Box plot illustrating the distribution of T1-weighted hypointensities in patients with no optic nerve involvement (0), unilateral optic nerve involvement (1), and bilateral optic nerve involvement (2). (B): Scatter plot demonstrating the positive correlation between optic nerve lesion volume assessed on DIR sequence and the total volume of T1-weighted hypointensities, adjusted for total intracranial volume (TIV).

General and regional brain volume analysis revealed multiple important findings in patients with optic nerve involvement. There was a statistically significant association between ONLV and cerebral white matter atrophy. Specifically, higher ONLV correlated with a reduction in cerebral white matter volume (β =-107.02, p = 0.016). In contrast, no correlation was detected between the number of optic nerves affected and the atrophy of brain structures. The precise volumetric assessment of brain structures revealed overlapping patterns of atrophy across both 3D T1W and 3D FLAIR sequences, particularly in regions associated with the visual pathway and occipital lobe. Significant atrophy was observed in the cuneus, lingual gyrus, fusiform gyrus, pericalcarine cortex, inferior temporal gyrus, and lateral occipital cortex. Additional volume reductions were detected in the isthmus of the cingulate cortex and the precuneus. Atrophy was also present in subcortical structures, including the thalamus and hippocampus. Global volumetric measures, including cerebral white matter and total gray matter volumes by optic nerve lesion status, are summarized in **Table 3**, while full volumetric data across all segmented brain structures are available in **Supplementary Material 1.**

When analyzing association between the volume of WMH on FLAIR sequence and brain atrophy, significant relationship was observed between periventricular white matter lesion volume and global brain atrophy. This analysis was conducted across the entire study cohort, including both patients with and without ONLV. Increased periventricular lesion burden was associated with a reduction in total brain volume (β = - 5058.08, p < 0.001), subcortical gray matter volume (β =-396.94, p < 0.001), cortical gray matter volume (β =-1426.82, p < 0.001), total gray matter volume (β =-2119.42, p < 0.001), and cerebral white matter volume (β =-2849.80, p < 0.001). At the same time, WMH localized in juxtacortical, deep white matter, and infratentorial regions did not show any impact on the atrophy of brain structures. These results are presented in Table 4.

**Table 4.** Correlation between WMH lesion volume, categorized by localization, and brain volume metrics in MS patients.

| Lesion localization / brain structures | Brain | Subcortical GM | Cortical GM | Total GM | Cerebral WM |
| --- | --- | --- | --- | --- | --- |
| **Periventricular** | **-5058.08 (p=0.000)** | **-396.94 (p=0.000)** | **-1426.82 (p=0.000)** | **-2119.42 (p=0.000)** | **-2849.80 (p=0.000)** |
| **Deep WM** | 13132.68 (p=0.335) | 536.57 (p=0.451) | 4890.87 (p=0.394) | 4349.12 (p=0.541) | 9471.13 (p=0.205) |
| **Juxtacortical** | 2766.74 (p=0.593) | 389.00 (p=0.151) | 124.08 (p=0.955) | 618.71 (p=0.819) | 2226.22 (p=0.432) |
| **Infratentorial** | -31092.34 (p=0.146) | -1034.29 (p=0.355) | -15732.23 (p=0.081) | -19384.47 (p=0.083) | -10272.91 (p=0.381) |
| **Optic nerves lesion volume** | -145.59 (p=0.071) | -0.64 (p=0.879) | -32.92 (p=0.331) | -33.16 (p=0.429) | **-107.02 (p=0.016)** |

Abbreviations: WM – White Matter; GM – Gray Matter.

Additionally, the analysis of T1W hypointensities in patients with ONLV revealed a significant association with greater atrophy across all 101 analyzed brain structures in the T1W sequence, even after correction for TIV (p < 0.05).

## Discussion

The primary objective of this study was to assess whether ONLV, as detected and segmented using the DIR sequence, corresponds with a more severe radiological profile in MS. Specifically, we evaluated its association with WMHs on 3D FLAIR, T1W hypointensities (black holes), and both global and regional brain atrophy. By comparing MS patients with ONLV to those without, we aimed to determine whether optic nerve pathology serves as a marker of widespread neurodegeneration and increased disease severity, as reflected in MRI findings. This study is among the first to quantify ONLV in the DIR sequence, providing a precise evaluation of how localized optic nerve demyelination relates to WMH burden, T1W hypointensities, and broader neurodegeneration processes in MS, potentially refining imaging-based biomarkers for disease severity and prognosis.

### ONLV and WMHs

Our study identified a strong correlation between ONLV (as detected in DIR) and periventricular, deep white matter, and juxtacortical WMH burden, with the strongest association observed for periventricular plaques. To our knowledge, this is the first study to systematically assess the relationship between the ONLV and the volumes of WMHs segmented by anatomical localization. While it is well established that lesions in the optic radiations are associated with retinal nerve fiber layer thinning and prolonged visual evoked potentials (VEPs) (Gabilondo et al., 2014; Sinnecker et al., 2015), no prior work has comprehensively investigated whether a similar volumetric relationship exists between optic nerve lesions and WMH burden across specific brain regions.

Notably, previous studies have demonstrated that optic nerve lesions may often be detected in asymptomatic patients, even in those with normal VEPs or no clinical history of ON, highlighting the importance of sensitive MRI techniques such as DIR (Davion et al., 2023; Hadhoum et al., 2016; London et al., 2019). Our findings extend this body of research by suggesting that optic nerve lesions are not isolated events but may reflect a broader pattern of disease dissemination, correlating with more extensive white matter involvement. Given that higher WMH burden has been associated with a more aggressive disease course and increased disability, ONLV detected via DIR could potentially serve as a novel biomarker for identifying patients at risk for more severe MS (Filippi et al., 2020; Gabilondo et al., 2014; Reich et al., 2009).

### ONLV and T1W hypointensities (black holes)

We observed a significant relationship between ONLV and T1W hypointensities (black holes). Patients with bilateral optic nerve involvement exhibited a significantly higher total volume of T1W hypointensities within the white matter compared to those with unilateral or no optic nerve lesions. Additionally, a direct positive correlation was observed between ONLV and T1W hypointensities (p < 0.001), indicating that greater ONLV is associated with more extensive irreversible white matter damage.

To our knowledge, no previous studies have investigated the relationship between optic nerve lesion volume and the volume of T1-weighted hypointensities. The only partially comparable work is that of Gabilondo et al., who manually segmented T1W hypointensities in the optic radiations and found that their burden correlated with both OCT-derived retinal thinning and volumetric atrophy of the visual cortex (Gabilondo et al., 2014). However, a direct comparison of ONLV and white matter black holes has not yet been reported in the literature.

T1W hypointensities are well-established imaging markers of disease severity and progression in MS, reflecting axonal loss and irreversible neurodegeneration (Calvi et al., 2022; Filippi et al., 2021, 2020). Our study demonstrated that T1W hypointensities directly contribute to greater atrophy across all 101 analyzed brain structures, which is in line with findings reported by other authors (Rocca et al., 2015). The greater severity of T1W hypointensities observed in patients with ONLV suggests that their presence may exacerbate widespread neurodegeneration in this subgroup. These findings highlight the clinical value of combining ONLV with T1W hypointensities assessment as an integrated biomarker for monitoring disease progression and identifying patients at risk of a more aggressive MS course.

### ONLV and atrophy

Our quantitative analysis revealed that higher ONLV was linked to a greater reduction in cerebral white matter volume (β =-107.02, p = 0.016). Furthermore, patients with ONLV exhibited significant atrophy in key regions of the visual pathway, including the cuneus, lingual gyrus, fusiform gyrus, pericalcarine cortex, inferior temporal gyrus, and lateral occipital cortex. Additionally, volumetric reductions were noted in the isthmus cingulate cortex and precuneus, which are involved in visuospatial integration and connectivity between visual and associative brain regions. This atrophy extended beyond the cortical occipital lobe into subcortical structures critical to the visual pathway such as the thalamus and hippocampus.

These findings align with the concept of trans-synaptic degeneration, where damage within the optic nerves propagates neurodegenerative processes throughout the brain. This mechanism extends beyond localized lesions and affects wider neural networks. Previous studies have reported cortical thinning in the occipital lobe of MS patients, particularly in those with a history of ON, as well as alterations in higher-order associative regions, findings that are consistent with our analysis (Eshaghi et al., 2017; Fisniku et al., 2008; Giorgio et al., 2008). Additionally, thalamic atrophy has been identified as a strong predictor of cognitive impairment in MS, with severe neurodegeneration serving as an early marker of disease progression (Azevedo et al., 2018; Eshaghi et al., 2017; Filippi et al., 2020; Fisniku et al., 2008).

### Periventricular WMHs and general atrophy

In addition to ONLV, our study found that periventricular WMHs (detected on 3D FLAIR) were significantly associated with greater cortical gray matter atrophy, subcortical gray matter atrophy, total brain atrophy, and white matter atrophy.

These findings reinforce the concept that MS-related demyelination and neurodegeneration are interconnected rather than isolated processes (Frischer et al., 2015; Kutzelnigg et al., 2005). The role of periventricular lesions as key contributors to neurodegeneration is well-established, as they disrupt major white matter tracts, including the corticospinal tract, corpus callosum, and associative fiber pathways (Fisniku et al., 2008; Jehna et al., 2015; Tóth et al., 2017). This damage leads to secondary neurodegeneration, further worsening atrophy in both cortical and subcortical brain regions. Thus, periventricular lesions represent an important marker for predicting long-term neurodegeneration in MS and could help identify patients at risk of a more aggressive disease course (Filippi et al., 2020).

Importantly, we observed consistent volumetric changes in both 3D T1W and 3D FLAIR sequences, aligning with prior studies that have compared MRI modalities for assessing MS-related brain atrophy (Goodkin et al., 2021). While 3D T1W is the standard for volumetric assessments, it is not always included in MS scanning protocols. Our findings demonstrate that 3D FLAIR-based volumetry may serve as a viable alternative in cases where 3D T1W-based assessments are unavailable or compromised.

### Clinical implications and future directions

The strong correlation between ONLV, WMHs, T1W hypointensities, and widespread brain atrophy suggests that optic nerve demyelination is a critical component of broader neurodegeneration. These findings also suggest that MRI-based ONLV may serve as a standalone marker that closely reflects other pathological changes captured by established MRI biomarkers. (Oertel et al., 2019; Petzold et al., 2017). As ONLV is strongly linked to the white matter degeneration and thus precise tracking could be crucial in modifying treatment strategies.

DMTs may be particularly beneficial in patients with early optic nerve involvement. If confirmed in longitudinal studies, MRI-based ONLV monitoring could become a key component of individualized MS treatment.

DIR has been demonstrated as the most sensitive MRI technique for detecting optic nerve lesions, even in patients without clinical symptoms or abnormalities in VEPs/ optical coherence tomography (OCT) (Hadhoum et al., 2016; Hodel et al., 2014). This highlights the importance of integrating DIR into standard MS imaging protocols. Additionally, DIR sequences offer high accuracy, ease of use, and reproducibility in detecting optic nerve lesions without requiring extensive neuroradiological experience (Sartoretti et al., 2017)

Future longitudinal studies should investigate ONLV in relation to other MRI-based biomarkers and clinical disability measures (e.g., EDSS scores, visual function tests) to better understand disease progression and treatment response. Further research assessing ONLV alongside WMH burden and T1W black holes in response to DMTs could refine therapeutic approaches, ultimately improving prognosis and quality of life in MS patients.

While this study provides valuable insights into the relationship between ONLV, white matter lesions, T1W black holes and brain atrophy in MS, several limitations must be acknowledged.

First, the cross-sectional nature of the study prevents us from tracking the longitudinal progression of disease-related neurodegeneration. A longitudinal approach would be necessary to assess how ONLV evolves over time and its direct impact on the other radiological MS findings. Additionally, higher uniformity and specificity of the study group would be crucial for evaluating the impact of the ONLV in longitudinal future study as well as correlation of ONLV with the clinical parameters. However, in our study, we aimed to compare patients with and without ONLV to investigate their direct impact on demyelination and neurodegeneration.

Second, manual segmentation of optic nerve lesions, though performed meticulously and validated by an experienced neuroradiologist, is a subject to inter-observer variability. The development of AI-based tools for automated segmentation could significantly enhance accuracy, objectivity, and reproducibility in future studies.

Also to provide a more comprehensive understanding of optic nerve pathology in MS, future research should incorporate complementary imaging techniques such as diffusion tensor imaging for microstructural analysis, OCT to assess retinal damage, and VEPs to evaluate functional impairment of the visual pathway.

This study focuses the role of ONLV and its association with WMHs burden, T1W blackholes and widespread brain atrophy in MS. Our findings indicate that ONLV correlates strongly with other established MRI biomarkers, highlighting its potential as a meaningful indicator of disease severity in MS. Future longitudinal studies incorporating automated volumetric assessments and multimodal imaging techniques combined with clinical parameters will be essential to further validate these findings and influence the treatment strategies for MS patients.

## Data Availability

All data produced in the present study are available upon reasonable request to the authors

