## Supplement Material 1 for "Optic Nerve Lesion Volume, White Matter Hyperintensities, and Brain Volumetrics in Multiple Sclerosis: A Multi-Sequence MRI-Based Analysis"

| Index | Variable | Range (mm$^3$) | | | Mean ± SD (mm$^3$) | | | Median (IQR; mm$^3$) | | |
|---|---|---|---|---|---|---|---|---|---|---|
| | | Zero nerves involved | One nerve involved | Two nerves involved | Zero nerves involved | One nerve involved | Two nerves involved | Zero nerves involved | One nerve involved | Two nerves involved |
| 1 | **X3Rd Ventricle** | 392.2–3662.0 | 418.2–3657.5 | 422.0–3875.1 | 1299.8 ± 509.0 | 981.1 ± 502.3 | 1625.7 ± 636.9 | 1359.5 (448.0) | 765.1 (444.5) | 1874.2 (770.1) |
| 2 | **X4Th Ventricle** | 1155.5–3785.1 | 1202.0–3413.2 | 1050.4–3896.3 | 2034.7 ± 480.3 | 1834.4 ± 443.3 | 2542.9 ± 709.7 | 1799.0 (697.7) | 1760.0 (484.3) | 2544.8 (1223.9) |
| 3 | **Brain Stem** | 14165.5–24994.5 | 15108.9–26539.6 | 11696.6–24861.6 | 21003.2 ± 2580.2 | 22342.1 ± 2840.1 | 19892.4 ± 2095.6 | 20313.5 (4805.8) | 23254.4 (4765.3) | 20542.6 (2552.1) |
| 4 | **Csf** | 257328.3–472337.3 | 245692.1–562936.8 | 246495.9–474533.0 | 359870.8 ± 42422.3 | 341925.8 ± 48355.8 | 363823.2 ± 48960.6 | 353252.8 (33022.3) | 355805.5 (56342.3) | 367784.8 (57387.7) |
| 5 | **Left Bankssts** | 1880.1–3290.0 | 1815.9–3101.8 | 1578.7–3486.2 | 2561.2 ± 413.5 | 2655.8 ± 324.0 | 2383.8 ± 298.2 | 2376.0 (721.6) | 2642.2 (516.6) | 2439.0 (363.2) |
| 6 | **Left Caudalanteriorcingulate** | 1962.3–4043.8 | 1839.3–3535.6 | 1740.2–3315.3 | 2607.1 ± 393.7 | 2742.3 ± 303.6 | 2440.0 ± 286.8 | 2518.0 (666.0) | 2878.5 (636.1) | 2431.3 (239.0) |
| 7 | **Left Caudalmiddlefrontal** | 4075.2–8627.8 | 3766.6–9310.6 | 3318.5–7608.8 | 6153.6 ± 1139.1 | 6768.9 ± 1404.5 | 5775.7 ± 796.9 | 6037.0 (1865.3) | 6653.8 (1610.3) | 5868.4 (635.3) |
| 8 | **Left Cuneus** | 3447.5–5079.1 | 2846.7–5025.4 | 2694.9–4549.3 | 4134.5 ± 498.1 | 4221.9 ± 523.9 | 3672.1 ± 417.2 | 3919.3 (1038.6) | 4296.3 (741.5) | 3529.5 (724.0) |
| 9 | **Left Entorightinal** | 2622.2–4409.3 | 2373.8–4036.0 | 2287.2–4068.4 | 3380.0 ± 408.6 | 3477.3 ± 363.6 | 3138.3 ± 301.2 | 3277.7 (723.8) | 3404.0 (349.1) | 3172.2 (392.4) |
| 10 | **Left Frontalpole** | 1949.6–3266.3 | 1933.5–2822.3 | 1734.4–3028.0 | 2515.0 ± 269.7 | 2522.3 ± 189.6 | 2382.8 ± 207.2 | 2441.6 (486.7) | 2546.0 (316.9) | 2395.2 (155.8) |
| 11 | **Left Fusiform** | 7827.8–12098.5 | 7402.3–11379.3 | 6907.3–11200.2 | 9551.8 ± 860.3 | 9918.3 ± 940.7 | 8955.7 ± 740.2 | 9329.8 (1555.3) | 9665.6 (1197.2) | 8871.3 (653.0) |
| 12 | **Left Inferiorparietal** | 7442.5–15427.8 | 6246.6–15348.7 | 5856.1–15439.2 | 10547.7 ± 1623.8 | 10940.0 ± 2561.5 | 9374.5 ± 1676.4 | 10185.8 (2542.0) | 10729.9 (3109.8) | 9445.8 (1484.5) |
| 13 | **Left Inferiortemporal** | 6315.3–11198.2 | 5039.7–10303.1 | 4910.6–11361.3 | 8365.6 ± 1343.7 | 8886.9 ± 1051.8 | 8088.7 ± 1128.9 | 8296.2 (1883.0) | 9238.0 (1706.6) | 8507.8 (1567.5) |
| 14 | **Left Insula** | 4185.0–7924.6 | 3701.2–6953.5 | 3453.6–7398.4 | 5722.3 ± 978.1 | 5947.3 ± 648.8 | 5298.1 ± 670.8 | 5492.0 (1619.9) | 6014.0 (918.3) | 5337.7 (487.5) |
| 15 | **Left Isthmuscingulate** | 2035.9–3884.5 | 1850.6–3105.9 | 1726.8–3581.7 | 2516.2 ± 347.4 | 2670.6 ± 293.4 | 2310.2 ± 268.4 | 2444.3 (345.6) | 2653.1 (533.1) | 2243.2 (223.7) |
| 16 | **Left Lateraloccipital** | 7005.1–13852.8 | 7176.5–12522.1 | 6763.8–12925.7 | 9920.2 ± 1248.1 | 10764.4 ± 1540.6 | 9320.9 ± 1168.2 | 9754.2 (1879.8) | 10750.2 (2977.4) | 9411.5 (1368.0) |
| 17 | **Left Lateralorbitofrontal** | 5225.0–9384.0 | 4027.2–8226.8 | 4047.7–8590.4 | 6742.0 ± 1197.2 | 7098.9 ± 1008.4 | 6158.0 ± 802.1 | 6442.4 (2008.1) | 7027.3 (1851.9) | 6262.0 (1077.9) |
| 18 | **Left Lingual** | 5633.6–9298.1 | 5196.0–8677.0 | 4873.8–8744.3 | 7315.4 ± 980.4 | 7597.8 ± 812.1 | 6551.6 ± 736.8 | 6844.3 (1004.1) | 7352.6 (1443.3) | 6321.1 (915.5) |
| 19 | **Left Medialorbitofrontal** | 3993.8–7259.4 | 3701.2–6267.7 | 3453.6–6332.4 | 5165.5 ± 644.7 | 5391.8 ± 685.9 | 4760.1 ± 496.2 | 4920.4 (897.9) | 5330.7 (1338.3) | 4738.2 (488.9) |
| 20 | **Left Middletemporal** | 6183.4–12073.3 | 4904.5–11174.8 | 4638.4–11351.6 | 8316.0 ± 1602.6 | 9604.6 ± 1299.0 | 8068.9 ± 1197.9 | 8573.1 (3376.3) | 9585.2 (2192.8) | 8279.7 (1424.1) |
| 21 | **Left Paracentral** | 2569.1–5015.9 | 2402.5–4274.7 | 1921.1–4818.5 | 3491.7 ± 716.5 | 3555.6 ± 475.2 | 3207.0 ± 553.1 | 3407.4 (1412.1) | 3635.1 (873.4) | 2962.2 (563.3) |
| 22 | **Left Parahippocampal** | 1967.3–3278.4 | 1850.7–3071.6 | 1726.8–2772.6 | 2528.1 ± 315.2 | 2554.5 ± 304.5 | 2315.5 ± 198.4 | 2438.6 (557.6) | 2521.3 (322.7) | 2362.2 (210.3) |
| 23 | **Left Parsopercularis** | 2785.0–7112.8 | 2538.4–4855.2 | 2548.2–5361.8 | 3810.5 ± 820.7 | 4349.2 ± 438.4 | 3823.5 ± 555.9 | 3943.6 (1401.6) | 4356.8 (635.6) | 3766.5 (491.2) |
| 24 | **Left Parsorbitalis** | 1741.5–3255.8 | 1460.9–2849.6 | 1292.6–2830.1 | 2260.2 ± 322.3 | 2410.0 ± 345.1 | 2049.5 ± 276.1 | 2146.6 (590.0) | 2375.4 (650.3) | 2089.6 (326.1) |
| 25 | **Left Parstriangularis** | 2995.3–5686.7 | 2536.8–4848.8 | 2022.1–5555.4 | 3769.4 ± 587.5 | 3945.3 ± 405.8 | 3547.0 ± 653.4 | 3822.5 (955.1) | 4158.7 (590.8) | 3320.1 (589.7) |
| 26 | **Left Pericalcarine** | 2372.1–4151.5 | 2428.2–3938.8 | 2276.9–4195.9 | 3418.1 ± 390.5 | 3450.1 ± 294.5 | 3019.0 ± 320.0 | 3310.6 (590.7) | 3509.0 (376.6) | 2927.5 (465.4) |
| 27 | **Left Postcentral** | 6729.8–12664.3 | 5827.5–12519.7 | 5335.6–11752.0 | 8829.5 ± 1372.7 | 9470.8 ± 1985.5 | 8124.2 ± 1199.6 | 8619.8 (2339.5) | 9784.7 (3015.4) | 7938.6 (1236.5) |
| 28 | **Left Posteriorcingulate** | 3881.1–7006.6 | 3597.0–6141.6 | 3311.5–6253.6 | 5146.1 ± 741.3 | 5149.3 ± 486.1 | 4669.5 ± 540.0 | 4916.3 (1299.3) | 5238.9 (936.5) | 4451.4 (506.9) |
| 29 | **Left Precentral** | 7345.8–14574.4 | 6657.7–13246.4 | 6101.5–12879.3 | 10208.2 ± 2001.3 | 10601.2 ± 1684.8 | 9527.9 ± 1337.4 | 10114.9 (4004.9) | 10945.6 (2586.8) | 9463.7 (1269.7) |
| 30 | **Left Precuneus** | 5807.3–10372.5 | 4463.5–10156.4 | 4388.8–11059.9 | 7642.0 ± 1256.8 | 8280.1 ± 1115.6 | 6957.5 ± 1043.2 | 7578.2 (1795.8) | 8159.8 (1751.0) | 6871.8 (1121.4) |
| 31 | **Left Rostralanteriorcingulate** | 1708.8–3856.5 | 1300.8–3418.8 | 1300.7–3685.7 | 2344.6 ± 455.6 | 2488.7 ± 279.5 | 2174.2 ± 317.5 | 2304.2 (723.6) | 2552.1 (486.0) | 2077.7 (273.4) |
| 32 | **Left Rostralmiddlefrontal** | 8294.5–17892.6 | 7668.5–15714.3 | 7369.6–18276.3 | 12370.8 ± 2590.8 | 12941.7 ± 1600.1 | 11197.5 ± 1665.6 | 11736.8 (3684.7) | 13561.8 (2229.5) | 11108.2 (2013.6) |
| 33 | **Left Superiorfrontal** | 12025.3–23294.4 | 10028.7–21110.7 | 10109.7–22048.5 | 16784.4 ± 3200.1 | 17523.0 ± 2444.3 | 14599.6 ± 2227.2 | 15505.1 (5470.7) | 17869.7 (3157.3) | 13687.0 (2494.4) |
| 34 | **Left Superiorparietal** | 7674.8–14699.4 | 6402.3–14367.7 | 6170.4–13569.1 | 10353.8 ± 1838.8 | 10964.4 ± 1386.2 | 9529.5 ± 1280.8 | 10273.1 (3973.7) | 11108.5 (1576.2) | 9859.4 (1533.0) |
| 35 | **Left Superiortemporal** | 6541.7–12971.1 | 4771.5–11973.2 | 4654.7–12222.6 | 8663.0 ± 1924.6 | 9201.8 ± 1552.2 | 7833.1 ± 1120.5 | 8293.0 (3789.6) | 9011.1 (1459.1) | 7948.7 (1006.1) |
| 36 | **Left Supramarginal** | 6299.7–12627.3 | 4845.8–11099.2 | 4651.1–11463.0 | 8931.4 ± 1361.2 | 8556.4 ± 1118.4 | 7847.4 ± 1328.4 | 8319.3 (1551.9) | 9209.5 (2121.1) | 8061.3 (1684.2) |
| 37 | **Left Temporalpole** | 3186.2–5201.8 | 2984.6–4582.2 | 2956.7–4488.6 | 3943.5 ± 436.3 | 4007.8 ± 371.5 | 3774.2 ± 325.2 | 3825.4 (553.4) | 3963.9 (584.5) | 3823.3 (412.0) |
| 38 | **Left Transversetemporal** | 1130.6–2524.3 | 1013.1–2001.6 | 952.6–2010.3 | 1573.3 ± 313.7 | 1615.0 ± 246.8 | 1393.9 ± 193.0 | 1472.6 (572.1) | 1587.3 (344.1) | 1327.2 (166.7) |
| 39 | **Right Bankssts** | 2108.1–3484.6 | 1850.6–3867.9 | 1726.8–3232.7 | 2701.5 ± 358.3 | 3060.8 ± 614.5 | 2537.2 ± 327.4 | 2594.3 (558.3) | 2837.7 (1237.5) | 2635.0 (528.6) |
| 40 | **Right Caudalanteriorcingulate** | 2687.9–4455.2 | 2231.8–4207.5 | 2094.1–4539.0 | 3431.5 ± 547.0 | 3683.7 ± 392.1 | 3150.9 ± 390.9 | 3375.3 (1149.3) | 3696.3 (634.0) | 3067.8 (417.5) |
| 41 | **Right Caudalmiddlefrontal** | 4157.8–9267.0 | 3772.1–8718.8 | 2992.4–8230.3 | 6217.3 ± 1684.0 | 6749.1 ± 1323.3 | 5777.9 ± 835.7 | 5908.8 (3013.6) | 6918.1 (2327.2) | 5727.8 (702.7) |
| 42 | **Right Cuneus** | 2602.0–4636.9 | 2321.6–3912.2 | 1990.9–4010.5 | 3382.4 ± 454.6 | 3461.6 ± 337.1 | 2951.9 ± 414.0 | 3123.1 (751.1) | 3519.8 (375.5) | 2808.0 (535.4) |
| 43 | **Right Entorightinal** | 2970.4–4632.6 | 2988.6–4321.7 | 2755.3–4223.0 | 3621.2 ± 361.7 | 3774.5 ± 378.5 | 3452.2 ± 284.0 | 3609.3 (577.1) | 3740.6 (655.7) | 3395.6 (285.9) |
| 44 | **Right Frontalpole** | 2065.5–3569.1 | 2054.7–3382.1 | 1929.0–3245.1 | 2778.8 ± 304.4 | 2757.6 ± 328.1 | 2581.4 ± 255.5 | 2659.9 (378.6) | 2619.6 (410.3) | 2596.6 (190.9) |
| 45 | **Right Fusiform** | 8068.7–13760.1 | 7457.6–13300.9 | 6995.2–12003.8 | 10382.4 ± 1407.7 | 11124.5 ± 1499.8 | 9673.8 ± 919.1 | 10050.6 (2984.4) | 10709.2 (2613.8) | 9665.0 (839.7) |
| 46 | **Right Inferiorparietal** | 9671.8–22181.4 | 8167.4–18395.7 | 7736.3–18210.8 | 13398.7 ± 1873.6 | 13582.3 ± 1990.3 | 12211.7 ± 1863.5 | 12870.2 (2280.2) | 13942.4 (3135.5) | 12971.1 (1898.3) |
| 47 | **Right Inferiortemporal** | 8305.0–13248.1 | 7299.1–12095.5 | 6899.4–13054.7 | 10102.6 ± 1432.0 | 10442.0 ± 1207.2 | 9271.0 ± 932.1 | 9329.8 (2899.2) | 10825.9 (2186.6) | 9314.3 (824.2) |
| 48 | **Right Insula** | 4792.4–8579.8 | 4080.7–7609.3 | 3884.9–8156.2 | 6284.6 ± 1040.7 | 6553.4 ± 767.8 | 5851.5 ± 720.9 | 6115.8 (2120.6) | 6649.1 (1182.6) | 5916.0 (502.2) |
| 49 | **Right Isthmuscingulate** | 3117.3–5143.1 | 2867.1–4757.5 | 2627.2–4572.1 | 4054.1 ± 538.2 | 4192.9 ± 420.8 | 3674.0 ± 377.7 | 3803.0 (447.5) | 4059.9 (659.7) | 3653.4 (248.8) |
| 50 | **Right Lateraloccipital** | 6765.2–14066.2 | 6759.1–12868.6 | 6037.4–12650.4 | 10181.3 ± 1199.8 | 10473.8 ± 1588.6 | 8947.7 ± 1200.0 | 10211.9 (962.2) | 10192.3 (2297.3) | 8521.6 (1334.9) |
| 51 | **Right Lateralorbitofrontal** | 5190.4–9944.7 | 4383.9–8746.2 | 4278.0–9012.5 | 6913.1 ± 1289.5 | 7421.3 ± 998.9 | 6355.2 ± 826.6 | 6676.2 (2416.9) | 7166.0 (1553.6) | 6402.0 (1044.4) |
| 52 | **Right Lingual** | 6488.2–10750.8 | 6220.2–9988.0 | 5907.5–9938.5 | 8587.8 ± 1012.0 | 8673.2 ± 895.5 | 7883.1 ± 779.3 | 8184.1 (1054.6) | 8814.4 (1322.0) | 7836.0 (916.6) |
| 53 | **Right Medialorbitofrontal** | 3978.4–7132.2 | 3701.1–6563.1 | 3453.6–6791.3 | 5230.5 ± 714.7 | 5539.8 ± 755.5 | 4815.5 ± 558.6 | 5063.0 (996.2) | 5502.3 (1403.1) | 4667.9 (478.3) |
| 54 | **Right Middletemporal** | 7352.4–12432.5 | 5517.4–11695.3 | 5506.5–12922.3 | 9455.0 ± 1750.0 | 9889.0 ± 1295.4 | 8759.4 ± 1296.1 | 8965.1 (3743.5) | 10243.0 (2133.7) | 9020.1 (1463.5) |
| 55 | **Right Paracentral** | 2606.6–5218.3 | 2609.2–5063.0 | 1894.8–5523.4 | 3730.9 ± 630.1 | 4023.1 ± 628.1 | 3528.0 ± 556.6 | 3795.1 (1281.5) | 3930.2 (805.1) | 3387.1 (554.2) |
| 56 | **Right Parahippocampal** | 2752.6–4374.2 | 2428.8–3986.9 | 2225.7–3819.3 | 3332.8 ± 395.5 | 3434.6 ± 323.8 | 3136.2 ± 306.1 | 3294.2 (681.1) | 3529.6 (539.3) | 3108.5 (276.6) |

| | | | | | | | | | | |
|---|---|---|---|---|---|---|---|---|---|---|
| 57 | **Right Parsopercularis** | 2730.6–5273.7 | 2447.2–5108.9 | 2451.9–5485.6 | 3636.0 ± 685.0 | 4175.9 ± 732.2 | 3497.2 ± 546.9 | 3749.6 (1226.4) | 3963.2 (1574.9) | 3309.0 (681.2) |
| 58 | **Right Parsorbitalis** | 3365.3–5250.4 | 3051.1–4977.8 | 2717.4–4703.0 | 4003.6 ± 398.6 | 4373.7 ± 487.1 | 3802.9 ± 394.1 | 3957.9 (785.7) | 4178.3 (958.0) | 3780.4 (493.2) |
| 59 | **Right Parstriangularis** | 4043.8–7244.6 | 3822.8–6284.2 | 3432.5–7425.6 | 5009.9 ± 503.4 | 5212.2 ± 469.3 | 4765.1 ± 559.8 | 4890.4 (752.4) | 5090.7 (652.7) | 4606.9 (509.7) |
| 60 | **Right Pericalcarine** | 2049.0–3659.9 | 1906.6–3314.9 | 1666.5–3260.3 | 2779.8 ± 391.5 | 2721.9 ± 250.0 | 2381.7 ± 278.0 | 2619.2 (664.5) | 2753.0 (318.4) | 2285.9 (206.8) |
| 61 | **Right Postcentral** | 6585.9–12742.8 | 5337.6–11375.9 | 5047.5–12601.8 | 8662.8 ± 1419.6 | 9196.7 ± 1582.8 | 8101.6 ± 1287.9 | 8257.6 (2682.2) | 9460.7 (2713.0) | 8055.0 (1078.5) |
| 62 | **Right Posteriorcingulate** | 3325.2–5340.7 | 2945.4–4984.3 | 2674.8–4886.6 | 4250.8 ± 529.0 | 4473.0 ± 503.7 | 3857.1 ± 434.8 | 4173.3 (1040.2) | 4537.3 (931.2) | 3741.9 (514.4) |
| 63 | **Right Precentral** | 8190.0–16544.2 | 7102.8–14030.2 | 6796.6–14364.7 | 11372.7 ± 1888.8 | 11470.0 ± 1564.1 | 10308.8 ± 1444.8 | 10839.1 (2991.0) | 11248.9 (2341.6) | 10009.8 (1690.7) |
| 64 | **Right Precuneus** | 7786.0–12243.4 | 6213.5–11349.0 | 6072.8–12517.2 | 9457.9 ± 850.3 | 9709.8 ± 782.5 | 8850.2 ± 839.4 | 9243.6 (937.5) | 9644.3 (1189.1) | 8797.7 (661.3) |
| 65 | **Right Rostralanteriorcingulate** | 1986.0–3640.9 | 1799.4–2710.4 | 1726.8–3851.4 | 2458.1 ± 321.7 | 2439.5 ± 169.0 | 2268.3 ± 243.0 | 2314.9 (429.5) | 2513.6 (305.7) | 2243.1 (173.5) |
| 66 | **Right Rostralmiddlefrontal** | 7992.6–19983.4 | 8373.0–15344.3 | 8002.3–20328.2 | 12525.2 ± 2028.8 | 12926.6 ± 1319.3 | 11439.4 ± 1669.2 | 12094.3 (4305.2) | 13531.4 (1817.8) | 11480.5 (1438.2) |
| 67 | **Right Superiorfrontal** | 11372.9–26291.2 | 9854.7–22846.4 | 10017.7–22739.0 | 17120.9 ± 3764.5 | 18265.1 ± 2958.9 | 15294.6 ± 2213.0 | 15927.5 (5711.2) | 18616.9 (3898.5) | 14883.6 (2079.0) |
| 68 | **Right Superiorparietal** | 8430.8–14833.3 | 7495.7–15957.5 | 7069.0–14541.6 | 11297.2 ± 1552.6 | 12414.6 ± 1900.9 | 10746.7 ± 1317.8 | 11388.8 (3152.5) | 12037.7 (1735.5) | 11049.7 (1387.2) |
| 69 | **Right Superiortemporal** | 6045.2–11914.6 | 5407.1–11463.3 | 5276.3–11625.5 | 8655.5 ± 2086.5 | 9319.3 ± 1243.5 | 8296.2 ± 1009.7 | 8926.3 (4079.4) | 9017.1 (1289.6) | 8642.8 (1117.2) |
| 70 | **Right Supramarginal** | 6820.1–11694.5 | 5839.8–10811.2 | 5114.4–11982.4 | 9090.4 ± 1597.4 | 9348.7 ± 1126.9 | 8214.1 ± 1206.8 | 8901.8 (3047.3) | 9474.5 (1867.4) | 7989.2 (1398.4) |
| 71 | **Right Temporalpole** | 2400.8–4834.1 | 2573.8–3886.1 | 2304.7–4029.2 | 3319.4 ± 497.3 | 3374.5 ± 342.3 | 3151.2 ± 324.0 | 3246.9 (889.7) | 3396.6 (597.5) | 3259.6 (428.2) |
| 72 | **Right Transversetemporal** | 1754.9–2687.9 | 1600.8–2585.2 | 1515.6–2459.7 | 2171.9 ± 266.9 | 2290.9 ± 202.3 | 2069.8 ± 178.0 | 2151.0 (586.9) | 2307.3 (402.0) | 2084.3 (230.1) |
| 73 | **Left Accumbens Area** | 393.0–825.1 | 341.9–847.5 | 379.7–829.7 | 613.9 ± 87.7 | 695.0 ± 81.6 | 608.3 ± 86.2 | 625.8 (150.7) | 727.0 (143.5) | 625.4 (83.9) |
| 74 | **Left Amygdala** | 1447.3–2176.8 | 1318.3–2025.8 | 1292.6–2157.8 | 1752.1 ± 173.4 | 1712.3 ± 178.6 | 1662.3 ± 177.4 | 1747.7 (314.7) | 1690.1 (358.2) | 1618.4 (202.2) |
| 75 | **Left Caudate** | 2941.3–4827.0 | 2202.2–5293.8 | 2427.6–5426.7 | 3793.0 ± 654.5 | 4351.7 ± 614.1 | 3623.8 ± 519.4 | 3835.7 (1375.7) | 4387.9 (861.4) | 3537.2 (656.1) |
| 76 | **Left Cerebellum Cortex** | 42709.7–66037.3 | 42118.0–60663.3 | 37978.0–63180.5 | 52207.0 ± 4935.2 | 53460.5 ± 5341.3 | 49764.2 ± 5364.5 | 52037.1 (7633.7) | 54434.1 (11979.1) | 50852.2 (7399.9) |
| 77 | **Left Cerebellum White Matter** | 10143.4–20050.1 | 13683.2–20898.6 | 11344.6–20331.6 | 15705.4 ± 2154.9 | 17379.0 ± 2171.7 | 15694.4 ± 1647.8 | 16050.9 (3962.1) | 17700.5 (2851.5) | 15764.0 (1223.7) |
| 78 | **Left Cerebral Cortex** | 205046.2–305034.1 | 180261.3–284201.5 | 188047.7–337990.3 | 239953.6 ± 27994.4 | 251147.2 ± 24518.3 | 232636.6 ± 21701.4 | 239115.8 (59223.4) | 260042.5 (42071.8) | 231960.8 (23986.8) |
| 79 | **Left Cerebral White Matter** | 173046.9–302842.8 | 143397.8–267463.6 | 137144.4–278735.2 | 220278.5 ± 32812.8 | 230702.2 ± 27185.6 | 202219.5 ± 23403.6 | 214578.2 (60398.4) | 227989.7 (44327.8) | 202399.1 (28761.2) |
| 80 | **Left Hippocampus** | 3700.4–5391.0 | 3194.2–5189.5 | 2347.5–5240.7 | 4205.6 ± 364.7 | 4305.7 ± 382.9 | 3930.2 ± 384.2 | 4337.7 (605.8) | 4260.4 (697.7) | 3817.6 (463.3) |
| 81 | **Left Inferior Lateral Ventricle** | 439.1–1905.8 | 461.2–1574.7 | 440.3–2826.4 | 717.0 ± 227.0 | 591.3 ± 162.1 | 830.0 ± 316.6 | 672.0 (128.3) | 546.7 (99.0) | 777.9 (387.3) |
| 82 | **Left Lateral Ventricle** | 3421.6–34923.4 | 3972.1–39774.3 | 3893.3–38732.4 | 10523.1 ± 6034.7 | 8362.7 ± 4685.7 | 14643.4 ± 6360.8 | 8402.7 (6191.1) | 7059.7 (1541.4) | 15966.2 (9234.0) |
| 83 | **Left Pallidum** | 1254.8–2238.3 | 1170.6–2025.8 | 795.3–1994.0 | 1672.1 ± 275.6 | 1735.2 ± 238.4 | 1483.2 ± 206.4 | 1610.3 (343.4) | 1834.7 (489.0) | 1465.4 (306.2) |
| 84 | **Left Putamen** | 4158.1–6738.0 | 3382.6–6595.3 | 3430.4–7390.8 | 5355.5 ± 750.8 | 5667.2 ± 492.3 | 5024.5 ± 587.1 | 5272.7 (1057.2) | 5639.8 (826.1) | 5014.1 (421.3) |
| 85 | **Left Thalamus** | 4664.9–9228.3 | 3563.5–8722.1 | 3604.3–10379.9 | 6892.2 ± 633.8 | 7350.7 ± 830.9 | 5925.6 ± 958.4 | 6851.4 (483.2) | 7691.0 (1065.8) | 5641.1 (1107.6) |
| 86 | **Left Ventral Dc** | 3150.4–5405.1 | 2784.6–4804.2 | 2517.2–5190.1 | 4267.5 ± 544.9 | 4229.3 ± 387.6 | 3691.2 ± 427.7 | 4077.5 (462.0) | 4354.0 (705.5) | 3616.6 (559.1) |
| 87 | **Right Accumbens Area** | 357.2–873.9 | 366.4–819.5 | 329.5–833.6 | 617.9 ± 77.2 | 690.7 ± 83.2 | 592.8 ± 86.4 | 621.8 (96.5) | 735.9 (116.5) | 616.3 (99.3) |
| 88 | **Right Amygdala** | 1482.8–2362.9 | 1439.9–2167.5 | 1376.6–2258.2 | 1890.1 ± 208.9 | 1793.1 ± 145.9 | 1707.2 ± 180.6 | 1832.3 (417.4) | 1753.9 (230.3) | 1653.9 (237.1) |
| 89 | **Right Caudate** | 3007.0–4980.6 | 2488.4–5298.6 | 2545.8–5526.0 | 3878.4 ± 661.3 | 4327.8 ± 579.4 | 3653.3 ± 536.7 | 3978.9 (1339.5) | 4465.4 (1124.9) | 3527.1 (789.4) |
| 90 | **Right Cerebellum Cortex** | 40038.9–66112.3 | 44119.4–61663.8 | 39805.4–61663.8 | 51639.4 ± 4741.4 | 53333.0 ± 5640.0 | 49785.1 ± 5331.5 | 51899.1 (7135.2) | 54286.2 (12068.5) | 51091.5 (7706.9) |
| 91 | **Right Cerebellum White Matter** | 11415.6–20254.0 | 13370.0–20390.1 | 11642.4–19807.9 | 15530.6 ± 2146.1 | 17036.0 ± 2190.9 | 15639.5 ± 1571.1 | 15899.4 (3982.9) | 17235.6 (3806.0) | 15864.1 (1779.5) |
| 92 | **Right Cerebral Cortex** | 205050.4–307429.2 | 181558.4–288497.3 | 186794.6–342221.4 | 241054.8 ± 28576.2 | 251936.6 ± 25530.3 | 233930.1 ± 22226.7 | 240728.2 (61881.3) | 259031.8 (39286.4) | 234247.8 (24828.5) |
| 93 | **Right Cerebral White Matter** | 170251.5–304899.6 | 146992.0–272484.6 | 140439.0–282026.8 | 220903.2 ± 33208.8 | 232121.4 ± 28673.5 | 203653.5 ± 23692.3 | 212625.9 (64503.3) | 229996.6 (44176.9) | 202350.9 (25052.3) |
| 94 | **Right Hippocampus** | 3351.1–5363.5 | 3293.4–5233.4 | 2567.9–5753.4 | 4421.9 ± 469.3 | 4375.7 ± 321.3 | 4001.8 ± 416.1 | 4433.5 (773.4) | 4413.6 (444.6) | 3897.3 (569.0) |
| 95 | **Right Inferior Lateral Ventricle** | 355.9–1846.3 | 391.8–1777.0 | 381.2–3121.5 | 722.4 ± 205.2 | 629.2 ± 184.5 | 808.0 ± 341.3 | 691.4 (138.5) | 599.3 (95.9) | 711.9 (399.9) |
| 96 | **Right Lateral Ventricle** | 3517.4–29867.7 | 4532.3–40959.4 | 3201.5–34923.3 | 8726.6 ± 4719.0 | 8293.6 ± 4464.3 | 13515.0 ± 5673.8 | 7322.5 (5494.2) | 8166.5 (2481.9) | 14512.1 (7718.6) |
| 97 | **Right Pallidum** | 1143.4–2184.2 | 1219.7–2110.6 | 1085.2–2006.1 | 1607.1 ± 288.0 | 1749.9 ± 196.1 | 1494.4 ± 194.2 | 1431.6 (378.2) | 1783.8 (327.2) | 1467.0 (282.8) |
| 98 | **Right Putamen** | 4242.3–6698.0 | 3472.0–6537.1 | 3480.1–6876.8 | 5156.5 ± 803.6 | 5552.8 ± 487.8 | 4822.2 ± 566.6 | 4836.7 (1120.7) | 5691.0 (660.7) | 4627.2 (604.6) |
| 99 | **Right Thalamus** | 4965.0–8643.1 | 4068.2–8215.6 | 3831.7–9577.5 | 6774.4 ± 696.8 | 6876.1 ± 787.6 | 5668.9 ± 854.3 | 6736.4 (588.3) | 7189.9 (1143.8) | 5363.7 (1247.1) |
| 100 | **Right Ventral Dc** | 2924.6–5052.1 | 3001.6–4777.2 | 2703.8–5143.1 | 4234.9 ± 451.6 | 4252.3 ± 371.9 | 3726.1 ± 406.0 | 4008.8 (719.3) | 4294.3 (565.8) | 3684.7 (562.6) |
| 101 | **Total Intracranial** | 1279871.0–1895828.6 | 1200203.0–1774819.9 | 1196157.0–1916333.0 | 1519303.5 ± 168346.3 | 1551741.5 ± 153122.3 | 1472619.1 ± 118884.5 | 1475535.2 (359825.3) | 1564180.5 (253288.0) | 1494956.9 (96497.4) |
